# Integrating Optometrists into Primary Health Care Facilities: Experience and Evidence from Sindh, Pakistan

**DOI:** 10.64898/2026.07.24.26358863

**Authors:** Khalid Iqbal Talpur, Fariha Sherwali, Waheed Ali Bhanbhro, Jalil Ahmed Rajper, Sadiq Bhanbhro

## Abstract

**Objective:** Visual impairment and preventable blindness continue to disproportionately affect rural communities in Pakistan, where the lack of trained eye care professionals at the Primary Health Care (PHC) level remains a critical gap in health service delivery. This study aimed to assess the contribution and practical implications of integrating Sindh Institute of Ophthalmology and Visual Sciences (SIOVS) trained optometrists at 95 Primary Health Care service delivery centres, known as Rural Health Centres (RHCs), across Sindh’s rural areas in 2025.

**Design:** Retrospective quantitative secondary analysis study.

**Setting:** Ninety-five Primary Health Care facilities commonly known as Rural Health Centres (RHCs) across rural districts of Sindh province, Pakistan.

**Sample:** We conducted a descriptive statistical analysis of 1,000 randomly selected patient records from participating RHCs.

**Results:** The analysis shows that optometrists independently managed 760 patients (76%), while 240 (24%) were referred to ophthalmologists at tertiary health care facilities for advanced care. Of the 118 cataract cases identified, all patients were successfully pre-registered for surgery and completed post-operative follow-up at their local RHC. Demographically, 55% of patients were aged 50 years or older, and women accounted for 52% of those examined, indicating that PHC facility-based optometry services improved access for groups traditionally marginalised in rural healthcare settings.

**Conclusions:** Our study demonstrates that an optometrist-embedded primary eye care model significantly reduces pressure on tertiary facilities, improves geographic access to essential vision services, and enables timely detection and referral of sight-threatening conditions. The analysis underscores the urgent need to scale up the deployment of qualified optometrists across all RHCs in Pakistan and calls for investment in diagnostic equipment and tele-ophthalmology infrastructure, as well as the integration of optometry into national PHC policy frameworks, to advance universal eye health coverage.

**STRENGTHS AND LIMITATIONS OF THIS STUDY:**

- The strengths of our study include our ability to analyse and report on patient records from first-ever interactions between optometrists and patients at the primary health care (PHC) level, assessing newly appointed optometrists’ contributions to eye care in PHC facilities across the 95 participating Rural Health Centres (RHCs) in Sindh province, Pakistan.
- This study is among the first in Pakistan to assess the integration of optometrists into RHCs using routine service delivery data.
- The use of a systematic random sample of 1,000 patient records, together with high inter-rater reliability (Cohen’s κ = 0.87), strengthened the consistency of data extraction and the validity of the findings.
- The study provides policy-relevant evidence to support strengthening primary eye care services by integrating optometrists at PHC level to improve access to eye care at PHC level in Pakistan.
- Limitations of this study include analysis of only patients’ records available at the facilities and maintained by the optometrists themselves, which may have introduced reporting bias. Including a qualitative component to interview patients and other RHC staff could have added value to this study and minimised potential bias.

## INTRODUCTION

Vision impairment is a major global public health challenge. It is estimated that 295 million people worldwide live with moderate-to-severe vision impairment, with uncorrected refractive error accounting for the largest single burden [1]. More broadly, the World Health Organisation (WHO) estimates that approximately 2.2 billion people globally live with vision impairment or blindness, of whom at least one billion cases could have been prevented or remain inadequately addressed [2]. Available evidence shows that uncorrected refractive error is the most common cause of avoidable vision impairment worldwide, and correcting it requires only a basic optometric assessment and the provision of spectacles, services well within the scope of a trained optometrist [1]. The challenge requires a shift from specialist-centred models of care to primary health care (PHC) integrated eye health systems, which need to strengthen primary eye care systems capable of delivering timely detection and intervention.

The burden of vision impairment falls disproportionately on low- and middle-income countries (LMICs), where over 90% of people with vision impairment reside despite having the lowest density of eye care specialists [2]. These patterns underscore the structural inadequacy of ophthalmologist-centred service delivery models in resource-constrained settings and highlight the essential role of allied eye care professionals, particularly optometrists, in delivering sustainable eye care. Visual impairment and blindness remain among the most pervasive yet preventable public health challenges facing LMICs. South Asia bears a disproportionate share of the global burden of vision impairment, with Pakistan, Bangladesh, and India collectively accounting for over 350 million people with uncorrected refractive error. Cataract, another leading cause of preventable blindness, further compounds this burden [3]. Without robust frontline screening and referral mechanisms, cases frequently progress to advanced visual loss before surgical intervention is sought [4]. These outcomes are not inevitable but reflect fragmented eye care systems in which the absence of primary-care providers delays diagnosis and treatment.

In response, the WHO’s Primary Health Care (PHC) framework has increasingly emphasised integrating allied health professionals, including optometrists, into first-contact care settings as a cost-effective strategy for expanding coverage and reducing specialist overload [2]. The framework emphasises task-sharing, which is the deliberate redistribution of clinical functions from specialists to appropriately trained mid-level providers and has demonstrated effectiveness across multiple health domains in low-resource settings [5]. Within eye care, optometrists occupy a pivotal role in this model. Trained to assess visual acuity, diagnose refractive conditions, detect anterior and posterior segment pathology, and triage patients requiring specialist referral, they provide the most efficient gateway to a well-functioning continuum of eye care [6]. In eye care, extensive evidence from sub-Saharan Africa and South Asia shows that optometrist-led or community eye health worker models can achieve outcomes comparable to specialist-delivered care for a defined range of conditions, including uncorrected refractive error and early cataract detection [8,9].

Pakistan reflects many of the challenges experienced across LMICs. In South Asia, Pakistan bears one of the highest burdens of blindness relative to its population size, with a national prevalence of blindness estimated at 0.9%, driven primarily by cataracts, uncorrected refractive error, glaucoma, and diabetic retinopathy [7]. The National Eye Survey (2023) further documented that cataract alone accounts for approximately 47% of bilateral blindness, followed by corneal opacity (12%), refractive error (9%), and glaucoma (7%). These figures reinforce the importance of early detection and referral, functions most efficiently delivered by optometrists embedded within community health infrastructure. Within Pakistan, the province of Sindh illustrates with particular clarity the structural inequities that shape eye health outcomes. While urban centres such as Hyderabad and Karachi host well-equipped ophthalmological facilities, the vast rural interior of the province remains profoundly underserved by trained eye care professionals. Consequently, millions of residents reach middle age and older adulthood without ever receiving a formal eye examination. The shortage of optometrists, combined with weak frontline screening and referral systems, means that treatable conditions such as cataract often progress to advanced visual loss before surgical intervention is sought. As a result, many patients are forced either to seek care at distant, congested tertiary hospitals or to forgo treatment altogether, perpetuating avoidable vision impairment and blindness across rural communities.

Community-based optometry programmes reduced the prevalence of unaddressed refractive error by 18–42 percentage points over 12–36 months [10]. In rural Bangladesh, deploying trained optometrists to Union Health and Family Welfare Centres was associated with a 38% reduction in first-level ophthalmologist consultations for refractive conditions, freeing specialist capacity for surgical and tertiary care [11]. Comparable reductions in avoidable specialist visits have been documented in Nepal, Sri Lanka, and rural India, lending strong cross-national validity to the task-sharing model [6].

### Optometry workforce and referral challenges in Pakistan

Pakistan’s optometry workforce is characterised by severe numerical shortages, geographic maldistribution, and regulatory fragmentation. Fewer than 3,500 registered optometrists for a population exceeding 230 million—a ratio of approximately 1:65,700, far below the WHO-recommended threshold of 1:10,000 [12]. The Higher Education Commission recognises eighteen universities offering BS Optometry programmes; however, graduates are disproportionately absorbed by urban private-sector employers, leaving rural public health facilities chronically understaffed [13].

Regulatory ambiguity is a principal structural barrier to deploying the optometry workforce in Pakistan [14]. The absence of a clearly defined legal scope of practice means that optometrists working in public facilities frequently encounter resistance from facility administrators who are uncertain about their mandate and lack the legal standing to prescribe, even within internationally recognised optometric parameters [14]. 61% of surveyed optometrists practised without formal job descriptions or institutional protocols, leading to inconsistencies in the quality-of-service delivery [15]. These barriers notwithstanding, SIOVS has demonstrated through its RHC deployment programme that institutional commitment can overcome systemic inertia, thereby establishing the proof of concept this study seeks to evaluate rigorously.

Cataract is the most surgically treatable cause of blindness in Pakistan and globally; yet access to cataract surgery remains profoundly inequitable between urban and rural populations [4]. A 2024 national survey estimated that fewer than 35% of cataract-blind individuals in rural Sindh had ever been informed about surgical options, and that transportation costs and unfamiliarity with distant facilities were the primary barriers to uptake of surgery [16]. Optometrists at RHCs are uniquely placed to address both barriers: by conducting systematic fundus and lens-opacity assessments in situ, they can identify surgical candidates early, provide pre-registration counselling, coordinate logistics, and conduct post-operative follow-up, a continuum of care that has demonstrably improved surgical outcomes in comparable settings [17,18].

### Gender, access, and eye health in rural settings

Gender inequity in access to eye care is a well-documented yet insufficiently addressed dimension of visual health disparities in South Asia. Globally, women are disproportionately affected by vision impairment, accounting for approximately 55% of the blind population despite comprising half of the global population [2]. In Pakistan, cultural norms that restrict women’s independent mobility, combined with the prioritisation of male household members’ healthcare expenditure, create structural barriers to women seeking eye care at distant facilities [19]. Studies from comparable settings in rural India and Bangladesh suggest that locating services within walking distance of the community, as RHC-based optometry achieves, can increase women’s utilisation of eye care services by 25–40% compared with facility-distant models [9,11]. This evidence positions local optometry deployment as both a health equity and a gender justice intervention.

### Tele-Ophthalmology and digital integration in eye care in rural areas

Tele-ophthalmology is a complementary strategy for extending specialist capacity to rural settings without requiring a specialist’s physical presence. An evaluation of an asynchronous tele-consultation model in Gilgit-Baltistan, in which optometrists transmitted retinal photographs and clinical data to remote ophthalmologists, achieved diagnostic concordance exceeding 85% for diabetic retinopathy and glaucoma suspects [20]. A review of emerging tele-optometry programmes in Pakistan identified connectivity, hardware costs, and the absence of regulatory frameworks as the primary barriers to scale-up [21]. Integrating tele-ophthalmology capacity into the existing SIOVS RHC network could substantially enhance frontline optometrists’ access to specialist support for complex cases without requiring patient travel.

The Sindh Institute of Ophthalmology and Visual Sciences (SIOVS) in Hyderabad has operationalised this framework through its Ophthalmic Community Services Department. SIOVS has deployed 95 trained optometrists to Rural Health Centres (RHCs) across 18 districts of Sindh, making it the sole institution in the province to have achieved the systematic integration of optometry professionals into public primary healthcare infrastructure at this scale. This deployment model aims to ensure that communities previously reliant on informal or no eye care can access diagnostic services, corrective prescriptions, and timely referrals within their own locality [22].

Despite growing advocacy for such models, rigorous evidence on their clinical effectiveness and system-level impact in the Pakistani context remains limited. Most existing studies in this domain have either been conducted in other low-income settings or lack disaggregated data on the specific contributions of optometrists compared with other health workers [23,8]. This gap limits policymakers’ capacity to design evidence-based workforce strategies and allocate resources appropriately. This study addresses this gap by systematically analysing service delivery data from optometrists deployed through SIOVS at rural RHCs in Sindh in 2025, to generate actionable evidence to scale primary-level eye care across the province and beyond.

The study objectives include:

- Assess the proportion of eye care cases managed independently by optometrists at RHCs versus those requiring specialist referral.
- Evaluate the contribution of optometrists in cataract case identification, surgical referral coordination, and post-operative follow-up.
- Identify operational challenges and systemic gaps in the existing model.

## MATERIALS AND METHODS

A retrospective descriptive study was conducted to evaluate the clinical roles, patient management outcomes, and referral patterns of optometrists deployed at Rural Health Centres (RHCs) across rural Sindh. This design was selected because it is appropriate for analysing routinely collected service delivery data to generate evidence on programme effectiveness in real-world operational settings [24]. The study was reported in accordance with the Strengthening the Reporting of Observational Studies in Epidemiology (STROBE) guidelines for observational research.

The study was undertaken within the SIOVS Ophthalmic Community Services Department, which delivers primary eye care through a network of government Rural Health Centres located in rural districts of Sindh province. These facilities serve populations ranging from approximately 25,000 to 100,000 people and are situated in districts characterised by limited ophthalmological infrastructure and long travel distances to tertiary eye care centres. At these facilities, SIOVS-trained optometrists functioned as the sole eye care providers, delivering scheduled eye care services and coordinating referrals to Taluka Headquarters Hospital (THQ) and District Headquarters Hospitals (DHQs), surgical facilities within the respective districts, and SIOVS Hyderabad when clinically indicated.

The study population comprised all patients examined by SIOVS-deployed optometrists at participating RHCs during 2025. A systematic random sample of 1,000 patient records was selected from the patient register using a fixed sampling interval based on the estimated annual patient volume. Records were eligible for inclusion if they contained complete demographic information (age and sex), a documented diagnosis or clinical impression, and a recorded management outcome indicating treatment at the RHC or referral. Records with missing primary outcome data were excluded. No personally identifiable information was extracted, and all records were anonymised before analysis.

Data were collected using a structured data extraction form. Variables extracted from each patient record included age, sex, presenting complaint, optometric diagnosis, clinical management (managed at the RHC or referred to an ophthalmologist), type of referral facility, and, for patients with cataract, pre-registration for surgery, surgical completion, and post-operative follow-up attendance. Data extraction was undertaken by two trained research assistants who were blinded to the study hypotheses. To assess data reliability, 10% of the sampled records were independently re-extracted, demonstrating strong inter-rater agreement (Cohen’s κ = 0.87).

Data were entered into and analysed using IBM SPSS Statistics version 26.0 (IBM Corp., Armonk, NY, USA). Descriptive statistics, including frequencies, percentages, means, and standard deviations, were calculated for all study variables. The distributions of clinical diagnoses, management outcomes, and referral patterns were summarised using descriptive statistics. Chi-square tests were performed to examine associations between categorical variables, including sex, referral status, age group, and cataract diagnosis. Statistical significance was defined as *p* < 0.05 for all analyses.

## RESULTS

### Demographic characteristics of study participants

Of the 1,000 patient records analysed, 520 (52.0%) were female, and 480 (48.0%) were male. The mean age of patients was 47.3 years (SD = 14.6 years). The 50-year-old and older age group constituted the largest proportion of patients (n = 550, 55.0%), followed by the 30–49-year-old group (n = 310, 31.0%) and the under-30 group (n = 140, 14.0%). These demographic characteristics are summarised in Table 1.

**Table 1:** Demographic Characteristics of Patients Examined by Optometrists at Rural Health Centres, Sindh 2025 (N = 1,000)

| Variable | Category | Frequency (n) | Percentage (%) |
| --- | --- | --- | --- |
| Sex | Female | 520 | 52.0 |
|  | Male | 480 | 48.0 |
| Age Group | < 30 years | 140 | 14.0 |
|  | 30–49 years | 310 | 31.0 |
|  | 50 years and above | 550 | 55.0 |
| Residence | Rural village | 834 | 83.4 |
|  | Peri-urban | 166 | 16.6 |

### Clinical management outcomes

Optometrists independently managed 760 of the 1,000 patients (76.0%) at the RHC level, prescribing corrective spectacles, providing counselling, and initiating treatment for minor anterior segment conditions without specialist input. The remaining 240 patients (24.0%) were referred to ophthalmologists at district hospitals or the SIOVS tertiary centre in Hyderabad for conditions requiring specialised assessment or surgical intervention. The distribution of primary diagnoses across the full sample is presented in Table 2.

**Table 2:** Distribution of Primary Ocular Diagnoses Among Sampled Patients (N = 1,000)

| Ocular condition | Frequency (n) | Percentage (%) | Management |
| --- | --- | --- | --- |
| Refractive Error (uncorrected) | 412 | 41.2 | Managed at RHC |
| Cataract (any stage) | 118 | 11.8 | Referred for surgery |
| Conjunctivitis / Anterior Segment | 158 | 15.8 | Managed at RHC |
| Presbyopia | 112 | 11.2 | Managed at RHC |
| Suspected Glaucoma | 58 | 5.8 | Referred |
| Diabetic Retinopathy (suspected) | 47 | 4.7 | Referred |
| Dry Eye / Other Anterior Segment | 55 | 5.5 | Managed at RHC |
| Other / Undifferentiated | 40 | 4.0 | Mixed |

### Cataract case management and referral outcomes

A total of 118 patients were diagnosed with cataract by the deployed optometrists, representing 11.8% of the sampled population. All 118 patients were pre-registered for cataract surgery through the SIOVS referral coordination pathway, a 100% pre-registration rate. Surgical procedures were subsequently completed for all referred patients at SIOVS Hyderabad or affiliated surgical sites. Post-operative follow-up was conducted at the local RHC for all 118 patients, with optometrists assessing uncorrected and best-corrected visual acuity, monitoring for wound complications, and providing spectacle prescriptions where required. No major post-operative complications requiring re-referral were recorded in the data. Cataract management outcomes are summarised in Table 3.

**Table 3:**
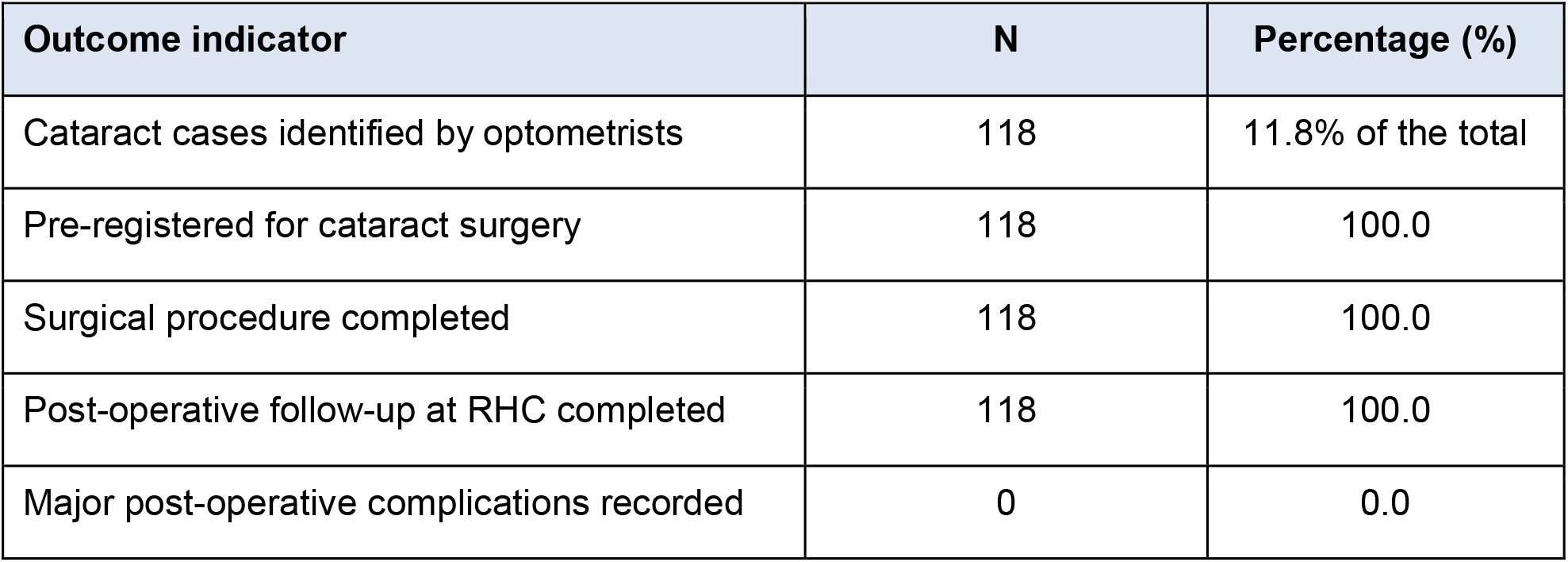
Cataract Case Management and Outcomes (N = 118)

### Association between age, sex, and clinical outcomes

Chi-square analysis revealed a statistically significant association between age group and cataract diagnosis (χ^2^ = 46.37, df = 2, p < 0.001), with 88.1% of cataract cases occurring in patients aged 50 years and above. No statistically significant association was found between sex and referral status (χ^2^ = 1.42, df = 1, p = 0.234), indicating that referral decisions were clinically driven rather than influenced by patient sex. The proportion of women utilising RHC-based optometry services (52%) exceeded the provincial female-to-male population ratio, suggesting that the availability of local services reduced traditional mobility barriers to accessing eye care for women.

## DISCUSSION

The findings of this study provide robust empirical support for the effectiveness of optometry-integrated primary eye care at Rural Health Centres in Sindh, Pakistan, and contribute to a growing body of international evidence on the value of task-sharing in low-resource eye health systems. The central finding that optometrists independently managed 76% of patients presenting at RHCs validates the clinical scope and utility of trained optometrists as first-contact providers in rural healthcare settings, consistent with evidence from comparable programmes in South Asia and sub-Saharan Africa [8,9,10].

The scale of independent case management documented in this study has direct implications for the efficiency of the healthcare system. In Pakistan’s predominantly specialist-centric eye care model, the absence of primary-level providers creates a cascade effect in which routine cases, such as refractive errors, presbyopia, and minor anterior segment conditions, are unnecessarily escalated to tertiary facilities, leading to overcrowding, long waiting times, and inefficient use of ophthalmologists’ time [14]. The SIOVS model inverts this dynamic: by resolving 76% of cases at the primary level, it frees ophthalmologists to focus on surgical and complex medical cases, optimising the entire care continuum. This finding aligns with WHO’s Integrated People-Centred Health Services framework, which emphasises that reorienting care towards the primary level is essential for achieving universal health coverage [2].

The cataract management data are particularly noteworthy. A 100% pre-registration, surgical completion, and post-operative follow-up rate among 118 cataract patients represents a standard of care coordination that is challenging to achieve even in well-resourced urban settings. In rural Pakistan, where cataract surgical uptake is estimated to be below 35% of identified cases due to logistical, financial, and informational barriers [16], this outcome reflects the transformative potential of optometrists functioning as care coordinators rather than merely as examiners. The optometrist’s dual role as clinical identifier and logistical facilitator addressed the classic ‘last mile’ challenge in cataract surgery programmes, ensuring that identification translated into intervention [17,18].

The demographic profile of patients served warrants careful interpretation. The predominance of patients aged 50 years and above (55%) aligns with the epidemiology of cataract, presbyopia, and age-related macular degeneration in Pakistan. It underscores the critical importance of accessible primary eye care for older adults [3]. Many of these individuals had likely never previously received a formal ocular examination, and the findings suggest that age-related conditions were being detected and managed for the first time through RHC-based services. This has important implications for planning: optometry services in RHC settings must be equipped and staffed to provide high-volume geriatric eye care, including fundus assessment for diabetic retinopathy and glaucoma screening.

The slight female majority among patients (52%) is a finding of particular policy relevance. Studies consistently document that women in rural Pakistan face greater structural barriers to accessing healthcare than men, due to mobility constraints, financial dependency, and prevailing cultural norms that deprioritise women’s health needs [19]. The fact that women utilised RHC-based optometry services at rates that approximated or slightly exceeded their population share suggests that local service delivery, which eliminated the need for long-distance travel to urban centres, meaningfully reduced these barriers. This aligns with international evidence that geographic proximity is the single most important determinant of healthcare utilisation among rural women in LMICs [9,11] and underscores the gender equity dimensions of primary care-integrated optometry.

The study also identified several operational challenges that temper an otherwise encouraging picture. Many RHCs lacked advanced diagnostic instruments such as slit-lamp biomicroscopes, non-contact tonometers, and digital fundus cameras, limiting optometrists’ ability to conduct comprehensive ocular health assessments and potentially resulting in under-detection of early glaucoma and diabetic retinopathy. The 5.8% and 4.7% rates of suspected glaucoma and diabetic retinopathy referrals, respectively, are lower than epidemiologically expected population rates, suggesting that detection may be constrained by equipment limitations rather than by the absence of disease [1]. Investment in diagnostic equipment and maintenance protocols is a prerequisite for the model to achieve its full potential.

Integrating tele-ophthalmology into the RHC optometry network is a high-priority opportunity for improvement. As demonstrated in Pakistan’s northern areas, asynchronous tele-consultation platforms enable optometrists to transmit retinal images and clinical data for specialist review, substantially improving diagnostic accuracy for posterior segment conditions without requiring patient travel [20]. Embedding this capacity within the SIOVS RHC network would extend the clinical reach of deployed optometrists, support their professional development through specialist feedback, and provide a scalable mechanism for managing the growing burden of diabetes-related eye disease in rural Sindh [21].

Finally, the model’s institutional sustainability warrants attention. SIOVS’s ability to deploy and sustain 100 optometrists at RHCs reflects extraordinary institutional capacity that cannot be assumed across all health system actors in Pakistan. Transitioning from an institutionally driven initiative to a government-mainstreamed programme, with optometry posts formally embedded within the provincial RHC establishment, under government salary scales and standardised job descriptions, is essential for long-term sustainability and national-scale replication [12,13].

## CONCLUSION

This study, based on a systematic analysis of 1,000 patient records, establishes that optometrists deployed at RHCs in Sindh, Pakistan, are effective, efficient, and indispensable frontline providers of primary eye care. By managing three in every four patients independently, coordinating 100% of identified cataract cases through to surgical completion and follow-up, and extending services to populations, especially older adults and women historically excluded from specialist-centric care models, SIOVS-deployed optometrists have demonstrated a compelling proof of concept for optometry-integrated primary healthcare in a low-resource setting.

The implications for Pakistan’s eye health policy are substantial. Universal expansion of this model, embedding trained optometrists in all RHCs and supported by appropriate diagnostic infrastructure, tele-ophthalmology connectivity, formal scope-of-practice legislation, and structured continuing professional development, could be the most impactful single intervention to reduce the burden of preventable blindness in Pakistan. Future research should evaluate the model using controlled study designs, longer follow-up periods, and quality-of-life outcome measures to build the evidence base required for national health policy adoption.

The SIOVS optometrist deployment model should be further strengthened and expanded to the remaining RHCs in Sindh province and considered for replication in other provinces as well, as an effective approach for improving access to primary eye care services in underserved communities.

## Abbreviations

PHC: Primary Health Care
RHC: Rural Health Centre
SIOVS: Sindh Institute of Ophthalmology and Visual Sciences
WHO: World Health Organisation
LMIC: Low- and Middle-Income Country
STROBE: Strengthening the Reporting of Observational Studies in Epidemiology
THQ: Taluka Headquarters Hospital
DHQ: District Headquarters Hospital

## FOOTNOTES

## Acknowledgements

The authors would like to thank the staff of the Sindh Institute of Ophthalmology and Visual Sciences (SIVOS), particularly the Ophthalmic Community Services Department, for their valuable support during data retrieval, management and analysis.

## Contributors

JAR conceived the idea for the study and identified the research topic. WAB developed the study design, led the research, analysed and interpreted the data, and drafted the manuscript. FS contributed to data collection and assisted with data analysis. KIT critically reviewed the manuscript and provided valuable intellectual feedback. SB supervised the study, provided academic guidance throughout the research process, critically revised the manuscript for important intellectual content, and approved the final version for publication. All authors contributed to the development of the manuscript, approved the final version, and accept responsibility for the integrity and accuracy of the work. SB is the guarantor of the study.

## Patient and public involvement

As this was a secondary analysis, patients and/or the public were not involved in the design, conduct or reporting of the study.

## Patient consent for publication

Not required.

## Ethics approval

Ethics approval was obtained from the Institutional Research and Ethics Committee of the Sindh Institute of Ophthalmology and Visual Sciences (Ref. No. SIOVS-IRB-2025-003). As the study involved analysis of anonymised retrospective records and did not involve direct patient contact, the requirement for individual informed consent was waived in accordance with institutional guidelines. Data confidentiality was maintained throughout the study, and no personal identifiers were retained in the analysis dataset.

## Data availability statement

Data are available upon reasonable request. We are happy to be contacted to share anonymised data. Please contact the corresponding author for requests.

